# Exploring influenza vaccination coverage and determinants through digital participatory surveillance

**DOI:** 10.1101/2024.11.08.24316973

**Authors:** Kathleen Kelley, Nicolò Gozzi, Mattia Mazzoli, Daniela Paolotti

## Abstract

**Background:** Vaccination is key for mitigating the impact of recurring seasonal influenza epidemics. Despite the efficacy and safety of influenza vaccines, achieving optimal vaccination uptake remains a challenge. This study aimed to explore the determinants of influenza vaccination uptake using data from Influweb, the Italian node of the InfluenzaNet participatory surveillance network, and to compare self-reported vaccination rates with data from official sources.

**Methods:** This study utilizes a longitudinal dataset of self-reported vaccination statuses from Italian participants across the 2011-2021 flu seasons. Logistic regression models were used to identify factors associated with vaccination uptake, while vaccination coverage of the Influweb population was compared with national data. Post-stratification weights were applied to account for demographic differences between the Influweb sample and the general population.

**Results:** The analysis reveals that individuals using public transport, those living with minors, and residents of the Islands macro-region are less likely to receive the influenza vaccination. On the other hand, university-educated individuals, and those on medication for chronic diseases are more likely to be vaccinated. Age also plays a role: individuals aged 44 and under are less likely to vaccinate compared to those aged 45-65, while those over 65 are more likely to do so. Furthermore, higher cumulative influenza-like illness incidence rates within a macro-region are associated with increased vaccination uptake, suggesting that local epidemic dynamics may influence individual decisions. Finally, the impact of COVID-19 pandemic was associated with an increase in influenza vaccination uptake. Comparison of the Influweb data to nationally reported vaccination rates revealed higher coverage for self-reported vaccination. This could be linked to the voluntary nature of the survey, possibly attracting a more health-conscious cohort.

**Conclusions:** Our study found that individuals living with minors and those relying on public transportation have lower odds of being vaccinated, despite having a higher documented risk of respiratory virus exposure. These findings highlight the importance of continued public health efforts targeting vulnerable groups and raising awareness about the risks of forgoing vaccination. The complex interplay of socioeconomic, demographic, and public health context significantly shapes vaccination decisions, emphasizing the need for tailored public health campaigns.

## Introduction

Vaccination stands as a fundamental pillar in public health, playing a key role in preventing infectious diseases and mitigating the impacts of epidemics and pandemics (1). Despite their proven efficacy and safety, vaccination programs often encounter challenges in achieving optimal uptake (2). This issue is particularly pressing in the case of influenza, where vaccine coverage remains below targeted rates despite the availability of effective vaccines (3). The World Health Organization (WHO) has recognized vaccine hesitancy—defined as delay in acceptance or outright refusal of vaccines despite available services—as a significant global health threat, underlining the need for targeted public health interventions to enhance vaccine acceptance (4).

It is well-established that a complex interplay of socioeconomic, demographic, and psychological factors contributes to vaccination uptake (5).

On a community level, economic factors and access to healthcare are significant determinants of vaccination rates. When vaccinations are easily available at pharmacies, workplaces, or community centers, the rate of uptake increases (6, 7).

The interaction of demographic and psychological factors also play a crucial role. Perception of risk, as a result of demographic characteristics, influences decisions. This can be related to the perceived risk of contracting the flu or potential side effects from the vaccine itself (6). Older adults and individuals with chronic diseases are more likely to get vaccinated due to a higher perceived risk of severe influenza outcomes (7, 8). In particular, patients with chronic kidney or liver diseases are more likely to vaccinate due to a higher vulnerability to severe influenza infections (7, 8). Better health literacy among older and more educated populations further contributes to higher vaccination rates (9). Conversely, lower perceived risk of infection typically leads to lower vaccination rates (6). Additionally, social encouragement from family, friends, coworkers, and especially healthcare providers, plays a crucial role in promoting vaccination (7, 8, 10).

Recommendations from public health authorities also play a role. In most industrialized European countries, the population groups that tend to get vaccinated the most are those for which the vaccine is recommended, namely elderly and fragile individuals, to the point that only these are the categories for which most national data are available.

While understanding well-documented vaccine determinants is essential, capturing evolving vaccination behaviors requires timely data collection methods. In contrast to traditional surveillance methods, digital participatory surveillance systems have gained prominence for their ability to collect real-time data on public health behaviors, symptoms, and vaccination uptake directly from volunteer participants. These systems not only provide valuable insights into individual health behaviors but also offer a faster approach to monitoring disease spread compared to traditional methods.

Participatory surveillance systems have been increasingly utilized to monitor influenza-like illness (ILI) and associated health behaviors. For instance, InfluenzaNet is a Europe-wide network monitoring influenza and other respiratory diseases through participatory surveillance (11). It has been used to identify key determinants associated with higher ILI risk, leveraging self-reported data to improve disease tracking across countries (12). In North America, the participatory surveillance system *Flu Near You* has been employed to assess health-seeking behaviors in individuals with likely ILI cases (13). Similarly, Australia’s Flutracking system has demonstrated adaptability by monitoring both influenza and COVID-19 incidence, highlighting the versatility of these systems in estimating illness trends (14).

In this study, we aim to investigate the determinants of influenza vaccination in Italy using longitudinal data over multiple influenza seasons from Influweb, the Italian node of InfluenzaNet. Our objective is to evaluate the potential of these platforms in identifying key factors influencing vaccination uptake and providing timely insights to guide more targeted vaccine campaigns. This approach could be particularly beneficial for improving uptake among groups that traditionally exhibit lower vaccination rates.

Through Influweb we have access to individual data on influenza vaccination decisions over multiple influenza seasons from 2011 to 2021, as well as a range of individual socio-demographic and health-related information, including age, employment status, household composition, education level, and medication use for chronic conditions. Additionally, we complement this data with reported influenza incidence rates. Logistic regression models were employed to identify determinants of vaccination uptake.

The results indicate that vaccination coverage among Influweb participants showed an upward trend, with a notable increase starting in the 2019-2020 flu season. Self-reported rates were higher than national rates, possibly due to the voluntary nature of the survey attracting health-conscious participants. Lower vaccination rates were associated with public transport use, living with minors, and residency in the Islands, while higher education, chronic disease management, and being 65 or older correlated with higher vaccination rates. The season coinciding with the COVID-19 pandemic significantly increased vaccination likelihood across all groups.

In conclusion, this study contributes to the existing literature by leveraging longitudinal data from a participatory surveillance platform to examine vaccination behaviors across multiple flu seasons. It shows how self-reported data from platforms like Influweb can offer valuable insights into individual health decisions, supporting the development of more effective public health strategies aimed at increasing vaccination rates and preventing outbreaks of vaccine-preventable diseases such as influenza.

## Methods

### Dataset description

This study utilizes a unique dataset provided by Influweb, which operates as a participatory symptomatic surveillance survey within Italy and forms a part of InfluenzaNet—a survey network dedicated to monitoring influenza-like illnesses across Europe. The survey leverages voluntary participation, where individuals partake by first providing demographic and health background via an intake survey, which gathers information such as age, presence of chronic diseases, education level, vaccination status, and region of residence.

The system allows participants to update their intake surveys. Participants then receive weekly email reminders to fill out a symptom survey, detailing any symptoms experienced in the past week and the health behaviors undertaken in response. The symptom surveys are routinely used to identify possible influenza-like-illnesses (ILI) cases, by referring to the ILI case definition from the European Center for Disease Prevention and Control (ECDC) (15). This study utilizes data spanning from 2011 to 2021, focusing on the determinants of vaccination status as reported in the intake surveys. There are a total of 9,646 intake surveys from 4,450 participants that were considered for this study.

Table 1 presents the characteristics of the participants used in the final dataset for the study. To calculate the distribution of these characteristics, the most frequently occurring value for each participant was used in cases where individuals changed categories (e.g., moved from one age group to another). Such changes were rare, occurring in only 28 participants.

**Table 1:** Participant characteristics and their distributions in percentages.

| **Characteristic** | **Categories** | **Percentage (%)** |
| --- | --- | --- |
| **Age Class** | 0-17 | 11 |
|  | 18-44 | 45 |
|  | 45-64 | 34 |
|  | 65+ | 10 |
| **Sex** | Male | 57 |
|  | Female | 43 |
| **Macro-Region** | Centre | 13 |
|  | Islands | 5 |
|  | North-East | 23 |
|  | North-West | 48 |
|  | South | 11 |
| **Education Level** | High school or less | 43 |
|  | Currently a student | 12 |
|  | University degree | 45 |

As a participatory system, Influweb relies on a self-selected sample of volunteers, which can introduce selection bias. For instance, participants in the Influweb study may have a greater interest in health-related topics, potentially leading to other characteristics or behaviors that differ from the general population. However, methodologies such as post-stratification can be employed to adjust the sample to more accurately represent the target population (16). Despite the potential for sample biases, the utility of Influweb has been well-documented. Digital participatory surveillance systems, like Influweb, offer unique insights into disease trends that are not accessible through traditional surveillance methods (17). Participatory surveillance systems are sensitive in detecting trends and early outbreaks because data can be collected and analyzed at a faster rate than traditional systems, which often rely on delayed reporting from primary care facilities (18). For example, ILI forecasting is improved when using Influweb data in conjunction with the sentinel data originating from primary care facilities (19). This is also being done with similar digital participatory systems across Europe (20, 21).

### Post-Stratification

For our study of the Italian population, we stratified the sample by sex and 5-year age groups for each year included in the study. We calculated the proportion of participants in each stratum *k* within the final dataset 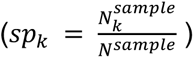 and compared it to the proportion of the Italian population in the same strata 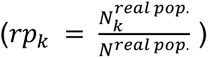. Population data was sourced from the Istituto Nazionale di Statistica, using the Intercensal Register for the years 2010-2019 and the Municipal Resident Population for the years 2020-2021 (22). Weights were then calculated by taking the ratio of the proportion in stratum *k* of the Italian population to the proportion of Influweb sample in stratum *k* for each year, using the formula: 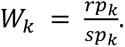

### Influweb Vaccination coverage

We assessed annual vaccination coverage from Influweb responses ranging from the 2011-2012 to the 2020-2021 flu seasons. A flu season spans from November 1 to May 1 of the following year, e.g., November 1, 2011 to May 1, 2012. Each participant’s data for a given flu season was represented by a single, unique intake form. In instances where multiple submissions were recorded for a participant within one season, the first one reporting vaccination was used. If no affirmative response was provided, the earliest intake form of the season was selected. The proportion of participants who reported vaccination out of the total participants for each season represents the vaccination coverage.

In addition to considering the overall vaccination coverage, coverage by macro-region and age class was considered. The Influweb data was then compared to the nationally reported data. National influenza incidence data was sourced from the Respivirnet report published by the Italian National Institute of Health (23). It reports ILI incidence at the regional level and is broken down into smaller age classes, so census data was used to adjust national figures to align with the survey’s broader age categories and macro-regions. Census data from 2011 was applied to flu seasons ranging from 2011-12 to 2018-19, while the 2019 census update was used for the 2019-20 season and subsequent years. This method ensured that the vaccination coverage estimates from the Influweb sample and the national population were comparable.

### Variable Description

Variables were selected based on their anticipated influence on vaccination status, guided by a literature review. These variables, listed in Table 2, include demographic, household, and health factors.

**Table 2:** Variables considered and their corresponding references.

| Variable | Values | Reference |
| --- | --- | --- |
| Sex | Male (reference), Female | Bertoni et al. 2022, Endrich, Blank, and Szucs 2009 |
| Public transport | True, False (reference) | L. Yang et al. 2017 |
| Lives with elders | True, False (reference) | L. Yang et al. 2017 |
| Lives with minors | True, False (reference) | Vaux et al. 2011, L. Yang et al. 2017 |
| Daily contacts with groups of 10+ people | True, False (reference) | Ibuka et al. 2016 |
| Education | High school or less (reference), Student, University | Barbadoro et al. 2013, Endrich, Blank, and Szucs 2009, Vaux et al. 2011 |
| Smoker status | True, False (reference) | Barbadoro et al. 2013 |
| Employment | Unemployed, Employed (reference), Retired | Barbadoro et al. 2013, Watkinson et al. 2023, Endrich, Blank, and Szucs 2009, Vaux et al. 2011, L. Yang et al. 2017 |
| Medication (proxy for chronic disease) | True, False (reference) | Barbadoro et al. 2013, Endrich, Blank, and Szucs 2009 |
| Age class | 0-17, 18-44, 45-64 (reference), 65+ | Barbadoro et al. 2013, Watkinson et al. 2023, Endrich, Blank, and Szucs 2009 |
| Pandemic (indicator that flu season is 2020-2021) | True, False (reference) | Watkinson et al. 2023, Y. Yang 2023, Leuchter et al. 2022, Porreca and Di Nicola 2023 |
| Macro-region dummy variables | Islands, Centre, North-west, North-east, South | Porreca and Di Nicola 2023 |
| Macro-region cumulative incidence | Numerical | Spruijt et al. 2016 |

Public transport indicates whether public transportation is the participant’s main mode of transport, in lieu of other options like bicycle or car. Household composition was assessed by determining whether participants lived with individuals in specific age groups. Participants who live with at least one person under 18 years old were classified as "living with minors," and those who live with at least one person aged 65 or older were classified as "living with elders." Both are binary variables.

Another variable was daily contacts with groups of people, where participants were asked if they regularly came into contact with groups of people (excluding those on public transport). If they reported daily interactions with groups such as more than 10 elderly individuals, patients, more than 10 children or teenagers (excluding their own children), or other groups of more than 10 people, the variable was coded as "True."

Educational attainment was classified into three levels. Participants who reported having a middle school or high school diploma were categorized as "High school or less," while those with a bachelor’s degree or higher were grouped under "University." Those still pursuing education were classified as "Students." Employment status was also categorized into three groups: "Employed" for individuals with full-time, part-time, or self-employment; "Unemployed" for students, homemakers, and those on long-term leave from work; and "Retired" for participants who reported that they were no longer working.

Participants who reported smoking either occasionally or daily were categorized as smokers, while those who did not smoke or were uncertain of their tobacco use were classified as non-smokers. Participants were coded as "True" for the medication variable if they reported taking medication for chronic conditions such as asthma, diabetes, chronic lung disease, cardiovascular disease, renal disease, or immunosuppression due to various therapies or conditions.

To assess for the influence of the COVID-19 pandemic, a binary pandemic variable was coded as "True" for data collected during the 2020-2021 flu season and "False" for prior seasons. Additionally, the cumulative incidence of influenza-like illness (ILI) was calculated per macro-region for each flu season. This continuous variable was derived from official regional case counts reported during each flu season.

A complete case approach was adopted for regression modeling of seasons 2011-2012 to 2020-2021. For variables of interest with significant missing values, imputation was employed using the latest available data from participants, which significantly reduced but did not eliminate missing data points. Most notably, the variable for education had 149 missing entries from 107 participants, and the macro-region variable had 78 missing entries from 56 participants.

The primary outcome, participant vaccination status, was initially categorized into three responses: yes, no, or unsure. Due to sparse data in the "unsure" category, with 58 entries from 33 participants, these responses were excluded from the final analysis.

The final dataset utilized consisted of 9646 responses from 4450 unique participants, represented by a single intake survey per participant for each flu season, as described for the vaccination coverage calculations above.

To assess the potential impact of the complete case approach, a sensitivity analysis was conducted by comparing vaccination coverage across seasons and the demographic breakdown of the sample before and after applying the complete case criteria. This analysis indicated that excluding cases with missing data did not significantly alter the vaccination coverage proportions across flu seasons or the demographics of the sample. These comparisons can be seen in Supplementary Figure S1 and Supplementary Table S1.

### Preliminary analysis

Statistical analyses began with individual chi-squared tests to examine associations between vaccination status and the levels of categorical covariates. The test compares the counts observed in each category of a data set to what we would expect to see if there was no relationship at all. If the differences between these observed and expected counts are large enough, the test suggests that the variables are related (24). These tests were used to identify potential factors that might influence vaccination uptake and for guiding the subsequent model selection process (25).

### Regression analysis

We consider a logistic regression model designed to estimate the odds of vaccination as a function of the identified significant covariates.

Logistic regression models are used to model binary outcomes. In this case, the outcome is whether a participant was vaccinated (1) or not (0). To facilitate the model selection process, the macro-region categorical variable was transformed into dummy variables. The initial full model included all variables identified from the literature review and the chi-squared tests.

The logistic regression equation can be expressed as:

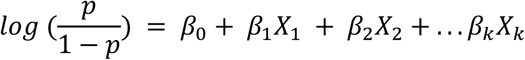

where *p* is the probability of vaccination, *β*_&_ is the intercept, and *β*_’_, *β*_(_, . . ., *β*_!_ are the coefficients for each corresponding predictor variable *X*_1_, *X*_2_, . . ., *X*_*k*_. For example, *β*_1_ might represent the effect of taking medication for a chronic disease (True/False), and *β*_2_ could represent the age category 65+ compared to the reference group 45-64.

#### Model Selection Procedures

To identify the most parsimonious model that adequately explains the data, we conducted two parallel model selection procedures starting from the full model:

1. “Drop-one” model selection: Initially, a "drop one" approach using likelihood ratio tests compared the full model against models each lacking one variable. This method systematically removes each predictor from a full model and compares the reduced model to the full model using likelihood ratio tests (LRT). The LRT assesses whether the reduced model (with one less covariate) fits the data significantly worse than the full model. The test statistic is calculated as:

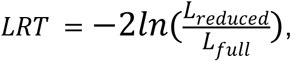

where *L*_*reduced*_ and *L*_*full*_ are the likelihoods of the reduced model and the full model. This statistic follows a chi-square distribution with degrees of freedom equal to the difference in the number of parameters between the full and reduced models (1 in this case). A variable is considered significant if its removal resulted in a significant LRT (p-value < 0.05), indicating that the model which excludes this variable fits the data significantly worse than the full model (26). Conversely, if the LRT yielded a p-value greater than 0.05, the variable was excluded because its removal did not significantly affect the model fit. By iteratively applying this procedure to all variables, we identified and retained only those predictors that significantly contributed to the model. This process resulted in a reduced model, referred to as the drop-one model.
2. Stepwise model selection: In parallel, a stepwise selection process was employed, considering both forward and backward selection. This procedure performed model selection by minimizing the Akaike Information Criterion (AIC) value. The AIC metric assesses a model’s likelihood while penalizing models with many covariates, defined as:

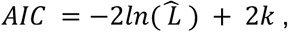

where *L̂* is the maximum likelihood of the model, and *k* is the number of parameters. The aim is to balance overall model fit with model complexity (27). Forward selection starts with no predictors, adding them one by one based on AIC improvement, while backward elimination starts with all candidate predictors from the full model, removing the least significant ones. The combined stepwise approach iterates between adding and removing covariates to achieve the lowest AIC, resulting in the stepwise selection model.

#### Model Comparison and Final Model Selection

After obtaining the drop-one model and the stepwise selection model, we compared them with each other and with the full model using likelihood ratio tests to determine the preferred model (28). The comparisons were as follows:

● Drop-One Model vs. Full Model
● Stepwise Selection Model vs. Full Model
● Drop-One Model vs. Stepwise Selection Model

The LRT was used to assess whether the simpler, nested model (with fewer covariates) provided an adequate fit compared to the more complex model. A non-significant p-value (p-value > 0.05) indicates that the simpler model is preferred due to its parsimony without a significant loss in model fit.

Based on these comparisons, the drop-one model was selected as the final model because it provided the best balance between model fit and simplicity.

#### Interpretation of the Final Model

The coefficients from the final, drop-one logistic regression model were exponentiated as *exp*(*β*) to obtain odds ratios (ORs), which quantify the association between each covariate and vaccination status (29). An OR greater than 1 suggests a positive association between the outcome and the covariate (higher odds of vaccination), while an OR less than 1 suggests a negative association (lower odds of vaccination).

## Results

### Vaccination coverage

#### Overall population

Figure 1 illustrates the annual vaccination uptake among the Influweb population (red) compared to the national population (blue), with data adjusted for age and sex. Throughout the earlier years of the study (2011-2012 through 2015-2016), vaccination rates among the Influweb participants were only marginally higher than those observed in the national data. Both populations show relatively stable vaccination rates during these years, with Influweb participants maintaining a vaccination rate in the range of approximately 20-30%, while the national rates hover slightly lower, typically between 10-20%. However, starting from the 2016-2017 flu season, there is a marked increase in the vaccination rate among the Influweb participants, which continues to rise steadily through to the 2020-2021 season. The national vaccination rates remain relatively stable with a smaller, though smaller, uptick in coverage for the last season.

**Figure 1:**
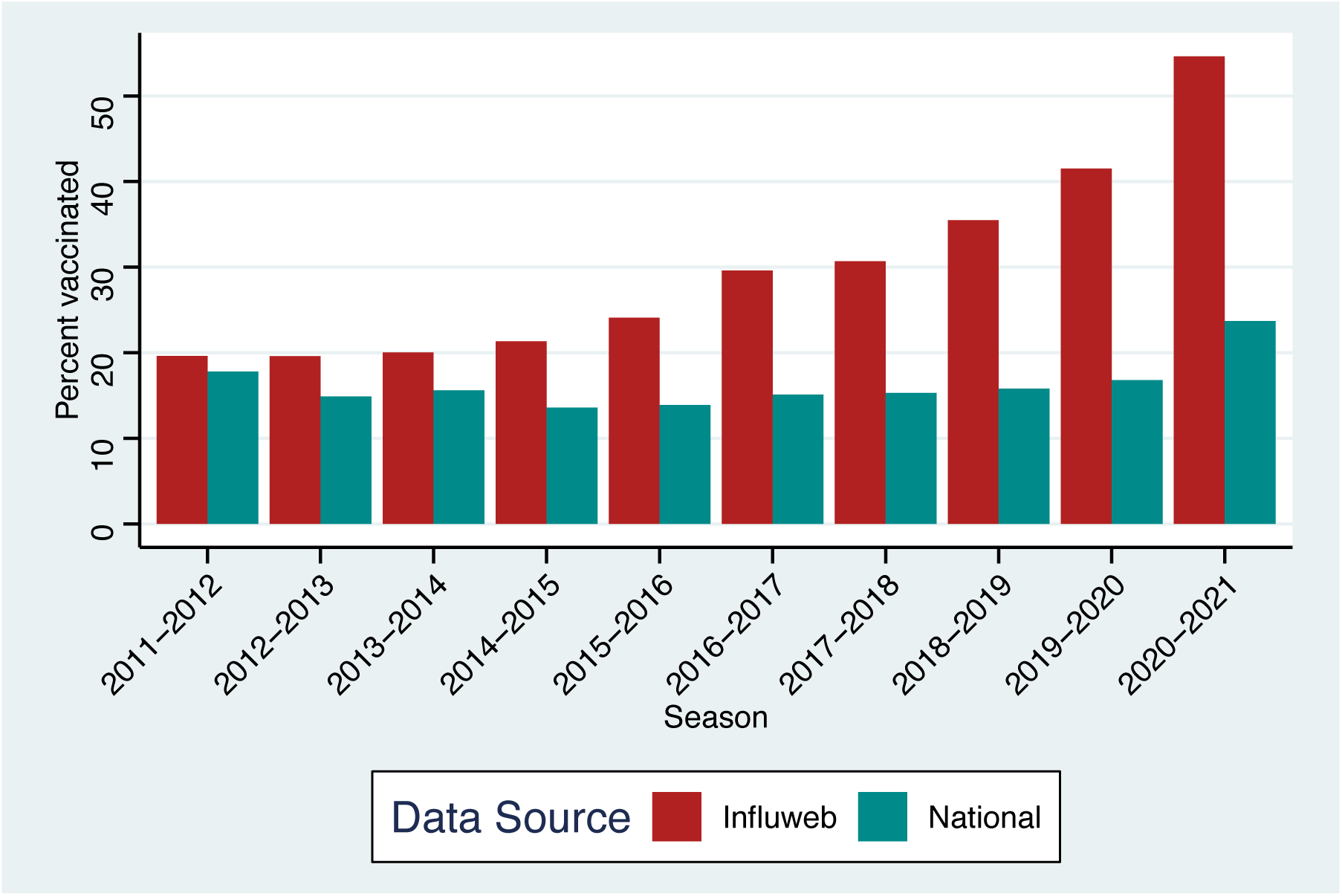
Annual vaccination uptakes reported by Influweb and national data, adjusted for age and sex

### Age-class and macro-region

We stratified our analysis on vaccination coverage in the Influweb population by age class and macro-regions and then compared it to the officially reported data. The 2011 and 2019 census data was utilized to align the survey population to the age groups and regions reported by the national data, allowing for comparison. It should be noted that the official data lacks a breakdown of vaccination coverage across the age groups below 65 years for the 2018-2019 flu season. This season still has vaccination coverage data broken down by regions like the others.

Figure 2 displays a comparison of self-reported vaccination rates from the Influweb study population versus vaccination rates from national data, broken down into four age groups: 0-18, 18-44, 45-64, and 65+. Each panel represents one of the age groups. Across all age groups, the red bars represent Influweb data, while the blue bars represent the national data.

**Figure 2:**
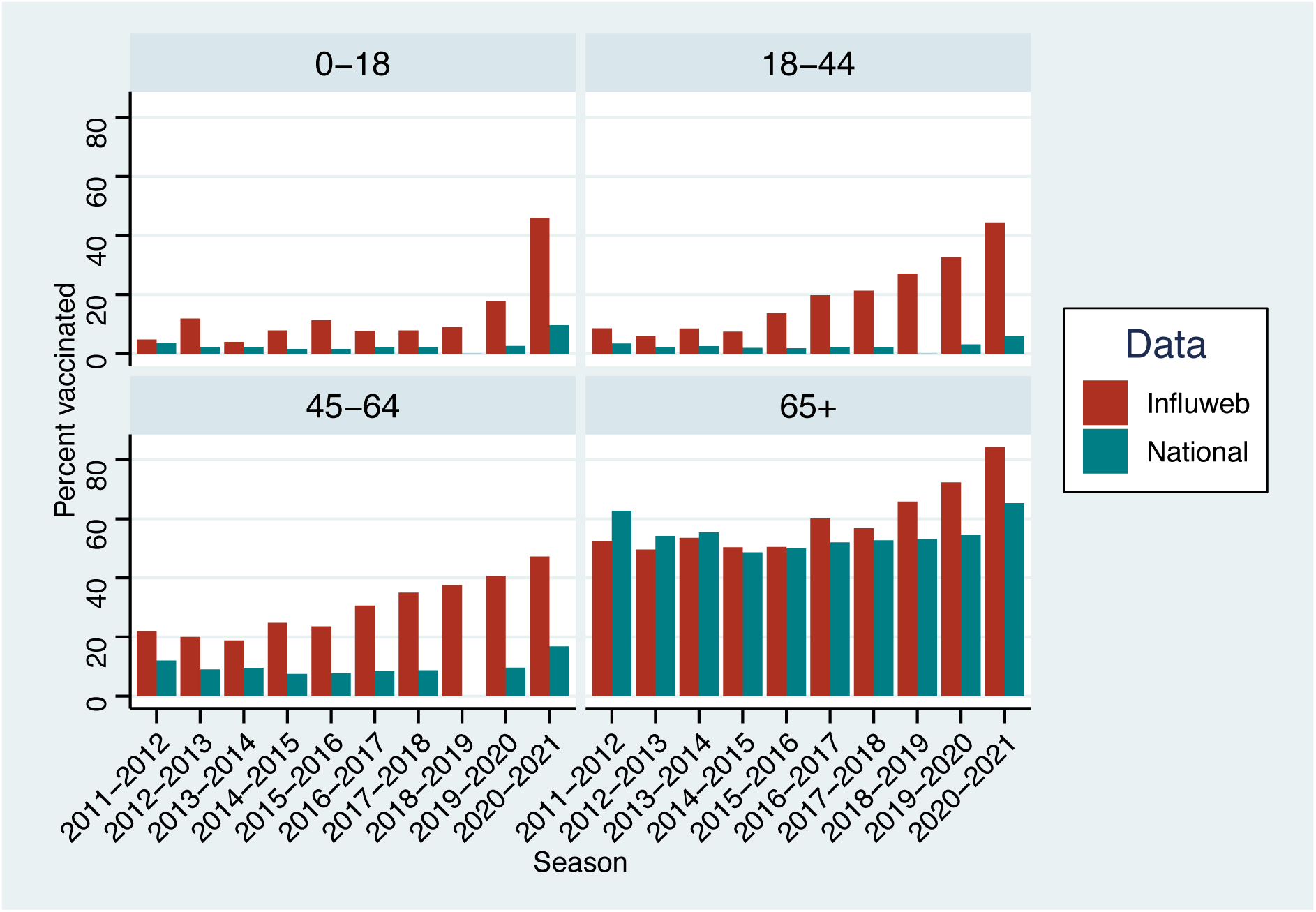
Comparison of vaccination rates from Influweb, adjusted for age and sex, and national data by age group

The vaccination rates for both the Influweb and national data in the 0-18 age group remain relatively flat across most seasons, with only slight fluctuations. However, there is a noticeable increase in vaccination rates for the Influweb population in the last two seasons, while the national rates remain low and steady.

In both the 18-44 and 45-64 age groups, the Influweb population shows a consistent upward trend in vaccination rates, particularly from the 2015-2016 flu season onward. During this period, a gap emerges between the Influweb and national data, with Influweb participants reporting increasingly higher vaccination rates, while the national data remains relatively stable.

In contrast, in the 65+ age group, both the Influweb and national data show an overall increase in vaccination rates over the years. Notably, the gap between the Influweb and national data in the later seasons is narrower in this age group compared to the younger age groups. This suggests that the Influweb’s 65+ age group has the highest similarity with national statistics among all the age classes.

Figure 3 illustrates the vaccination trends across the five geographical macro-regions in Italy. The graph compares self-reported vaccination rates from the Influweb study population (in red), adjusted for age and sex, to national data (in blue).

**Figure 3:**
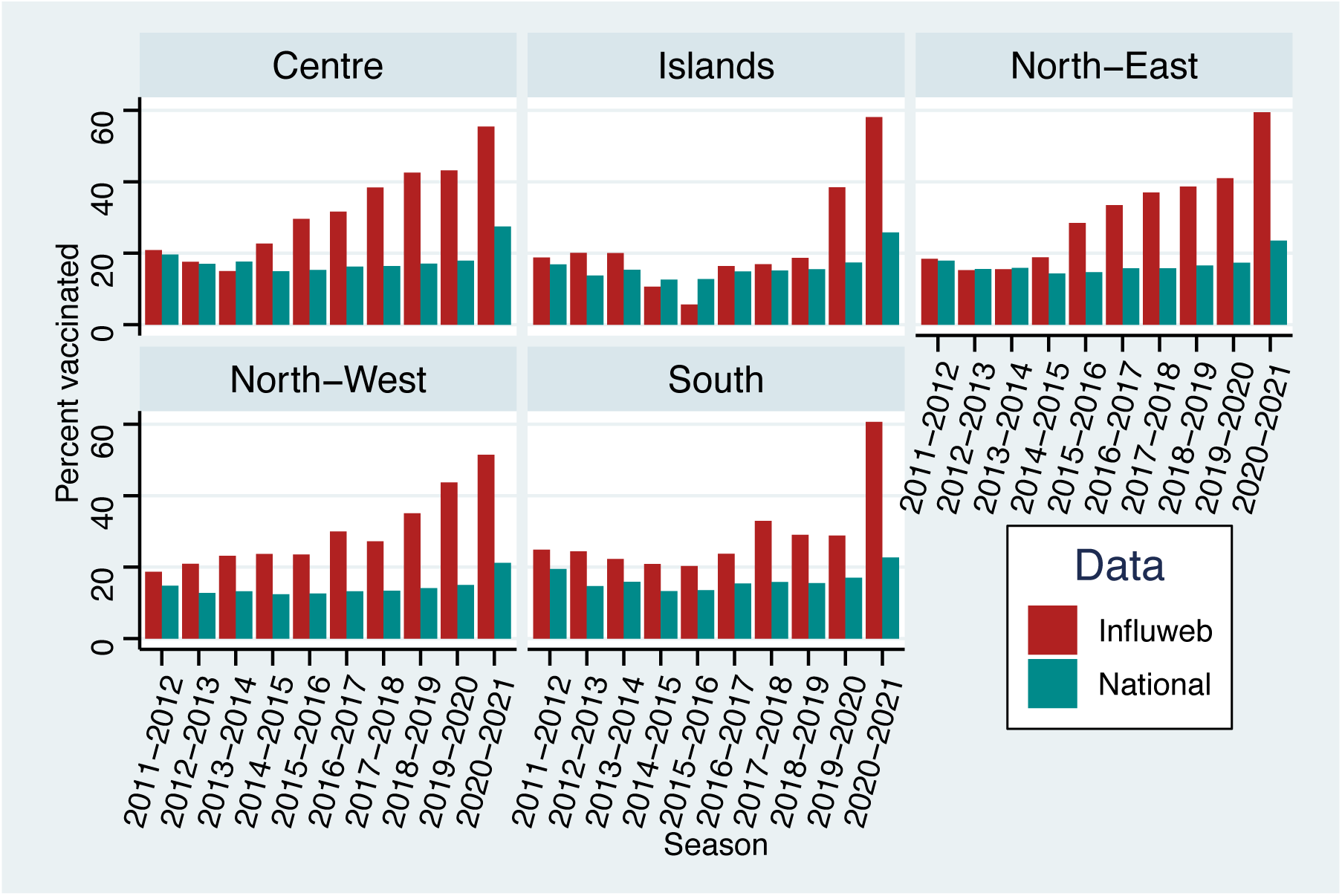
Comparison of vaccination rates from Influweb, adjusted for age and sex, and national data by macro-region

Throughout the years, the Influweb self-reported vaccination rates have been consistently higher than national rates in almost every season across all macro-regions. There is a general increase in vaccination rates over time, with a notable spike in the 2020-2021 flu season.

In the North-East and North-West regions, Influweb rates are relatively stable, except for the aforementioned increase in the last flu season. In contrast, the vaccination rates in the remaining regions exhibit more fluctuations. The proportion of Influweb participants from Islands, Centre and South macro-regions is smaller than that of the North-West and North-East macro-regions. As a result, the denominator for vaccination coverage within these macro-regions is smaller, and the vaccination rates are more sensitive to changes in participant numbers across seasons.

### Retrospective data comparison

To validate the consistency and accuracy of self-reported vaccination status within the Influweb population, we also explored retrospective vaccination data. In addition to inquiring about a participant’s vaccination status for the current flu season, participants were asked whether they were vaccinated in the prior season. The results indicate the retrospective vaccination rates reported for prior seasons closely matched the actual vaccination rates reported during those seasons, suggesting that the data are consistent over time. Both rates follow a similar trajectory with an upward trend over the seasons. For more details illustrating this comparison, please refer to the supplementary material and Supplementary Figure S2.

## Vaccination determinants

### Preliminary analysis

Table 3 shows the results of the chi-squared tests for categorical variables. The chi-squared tests indicated significant associations for all considered covariates except for Macro-Region. Due to these results, all covariates were retained for further exploration in the model selection process and the covariate “Macro-region” was broken down into dummy variables.

**Table 3:**
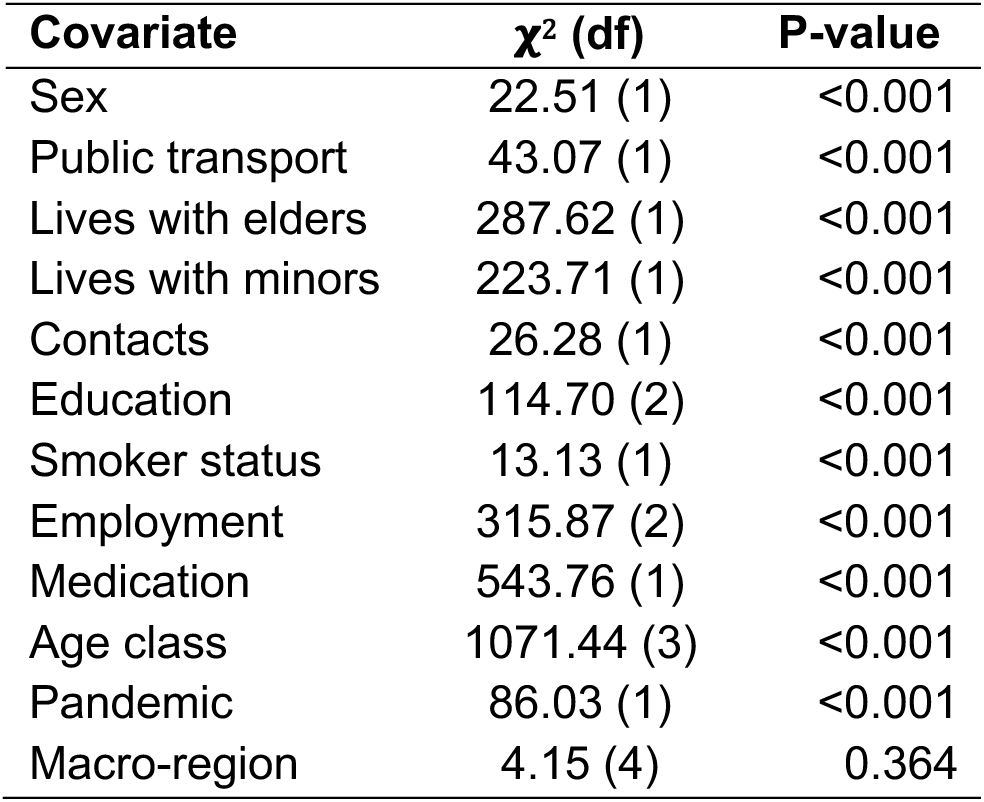
Chi-square test results.

### Logistic Regression Model

The logistic regression analysis aimed to identify significant predictors of influenza vaccination status, and the model demonstrated an overall accuracy of 77.1% (i.e., the percentage of samples that the model correctly classified). The results can be seen in Table 4.

**Table 4:** Logistic regression coefficients, odds ratios, and standard errors.

| **Variable** | **Odds Ratio** | **Coefficient** | **Std. Error** | **Significance** |
| --- | --- | --- | --- | --- |
| Public Transport | 0.776 | -0.254 | 0.090 | * |
| Lives with minors | 0.799 | -0.224 | 0.075 | * |
| Education: Student vs high school or less | 2.042 | 0.714 | 0.196 | * |
| Education: University vs high school or less | 1.220 | 0.199 | 0.071 | * |
| Employment: Retired vs Employed | 1.714 | 0.539 | 0.134 | * |
| Employment: Unemployed vs Employed | 0.958 | -0.043 | 0.093 |  |
| Medication | 2.217 | 0.796 | 0.072 | * |
| Age class: 0-18 vs 45-64 | 0.250 | -1.386 | 0.223 | * |
| Age class: 18-44 vs 45-64 | 0.575 | -0.554 | 0.070 | * |
| Age class: 65+ vs 45-64 | 2.002 | 0.694 | 0.131 | * |
| Pandemic | 7.675 | 2.038 | 0.161 | * |
| Islands | 0.45 | -0.798 | 0.204 | * |
| Macro-regional cumulative incidence | 1.265 | 0.235 | 0.029 | * |

Individuals who use public transport were found to have a reduced likelihood of being vaccinated against influenza, with the odds being 22.4% lower compared to those who use other methods of transportation (OR = 0.776). Similarly, living with minors was associated with a decreased likelihood of vaccination. Participants living with minors had 20.1% lower odds of being vaccinated compared to those who do not live with minors (OR = 0.799).

Education level also played a role in explaining vaccination status. Those with a university education had 22% higher odds of being vaccinated compared to those with a high school education or less (OR = 1.220). This indicates that higher educational attainment is positively associated with vaccination uptake. Students had more than double the odds of being vaccinated compared to individuals with a high school education or less (OR = 2.042), representing an increase of 104.2% in the odds of being vaccinated. Regarding employment status, there was no significant difference in vaccination likelihood between unemployed and employed individuals. Medication use emerged as a significant predictor, with individuals taking medication for chronic conditions having 121.7% higher odds of being vaccinated (OR = 2.217).

Age was another significant factor in vaccination decisions. Participants aged 0-18 had 75% lower odds of being vaccinated compared to those aged 45-64 (OR = 0.250). The 18-44 age group also exhibited a lower likelihood of vaccination, with the odds being 42.5% lower than the 45-64 age group (OR = 0.575). Conversely, those aged 65 and older had 100.2% higher odds to be vaccinated compared to the 45-64 age group (OR = 2.002).

The emergence of the COVID-19 pandemic was significantly associated with vaccination behavior. The analysis revealed that individuals had 667.5% higher odds of being vaccinated against influenza during the pandemic season (OR = 7.675). Geographic factors also played a role; individuals residing in the Islands macro-region had 55% lower odds of being vaccinated compared to those living in mainland regions (OR = 0.450). Additionally, higher macro-regional cumulative incidence was associated with increased vaccination likelihood. For each unit increase in regional cumulative incidence, individuals had 26.5% higher odds to be vaccinated (OR = 1.265).

### Reasons for vaccination

In addition to their demographic factors, Influweb participants who indicated they were vaccinated were asked to provide their reasoning for doing so, with the option to select multiple reasons from a given list. This can be seen in Table 5. The reasons provided by the participants align with some of the significant predictors identified in the regression model, offering additional context to understand their vaccination decisions.

**Table 5:** Reasons for vaccination among vaccinated participants.

| Reason for Vaccination | Percentage (%) |
| --- | --- |
| Vaccination decreases my risk of getting influenza | 52.74 |
| I always get the vaccine | 45.49 |
| I belong to a risk group (e.g., pregnant, over 65, underlying health condition, etc.) | 42.14 |
| Vaccination decreases the risk of spreading influenza to others | 32.55 |
| My doctor recommended it | 17.00 |
| I don't want to miss work/school | 15.68 |
| The vaccine was free (no cost) | 13.54 |
| The vaccine was readily available and vaccine administration was convenient | 11.50 |
| It was recommended in my workplace/school | 8.68 |
| Other reason(s) | 3.98 |

Over half of the participants (52.74%) reported that they chose to get vaccinated to decrease their personal risk of getting influenza, while 32.55% of participants cited reducing the risk of spreading influenza to others as a motivation. A significant percentage (45.49%) also reported that they always get the vaccine, possibly due to established health routines or enhanced access through retirement health plans. Moreover, 42.14% of participants indicated belonging to a high-risk group as a reason for vaccination.

## Discussion

The results of this study, conducted in Italy using data from the Influweb platform, provide a view of the factors influencing influenza vaccination uptake among the Influweb population. By examining vaccination coverage, logistic regression outcomes, and qualitative reasons for vaccination, these findings can be contextualized within the broader literature on vaccination behavior.

Our study found that those living with minors were less likely to vaccinate, which is contrary to findings from France (30) and Hong Kong (31), where living with children was associated with higher vaccination rates. It’s important to note that those living with minors are often at a higher risk of exposure to respiratory viruses, such as influenza and COVID-19, making this an important finding that warrants further investigation (32, 33). This discrepancy may stem from caregivers being inadequately informed about their actual risk of ILI, potentially leading to a false sense of security and lower vaccination rates. Targeted public health messages could be an effective way to raise caregivers’ perception of risk, helping it more closely reflect the reality.

Risk perception has been widely recognized as a key motivator in vaccination decisions, as demonstrated in multiple studies (7, 8, 34). In particular, these studies found that older adults and individuals managing chronic conditions experience a heightened perceived risk of severe influenza outcomes, making them more likely to get vaccinated. This is supported by our data. The logistic regression model revealed older age and taking medication for chronic disease to be strongly associated with vaccination likelihood, also aligning with research from Hong Kong (31) and Italy (35). Interestingly, while 42% of vaccinated participants cite belonging to a risk group as a motivating factor in their choice to vaccinate, only 17% note their doctor recommended vaccination to them. This, along with model results, implies the success of current public health campaigns informing those in high-risk groups of their increased risk from influenza, as well as a level of health literacy in the Influweb population.

Our logistic regression analysis revealed that Influweb participants who rely on public transportation as their main form of travel had 22.4% lower odds of being vaccinated. This finding aligns with Yang et. al (31), who highlighted transportation access as a key factor in vaccine uptake, with individuals who have private or more direct forms of transportation (e.g., a personal car or the ability to walk to a healthcare provider) being more likely to vaccinate. This is important because, like those living with minors, public transportation users are often at higher risk of exposure to respiratory viruses due to close contact with others in enclosed spaces (36). Public transportation may present barriers to vaccination access compared to more convenient options like walking or private transportation.

Vaccination convenience and accessibility plays a critical role in increasing uptake, as demonstrated by research from Abbas et. al (6) and Nagata et. al (7). These studies conclude that when vaccinations are easily accessible at pharmacies, workplaces, or community centers, the rate of uptake significantly increases. In our study, 14% of Influweb participants reported vaccinating because the vaccine was free, while 12% cited the convenience of availability and administration as key reasons for their decision. This underscores the importance of ensuring that vaccines are not only affordable but also easily accessible in locations that people frequent regularly.

Education level is another significant predictor of vaccination, with higher education correlating with increased vaccination rates. Participants with university-level education were more likely to vaccinate, consistent with the findings from China by Gong et. al (34) and France by Vaux et. al (30). Similarly, a study by Wang et. al (9) suggests that better health literacy and awareness of more educated populations drives higher vaccination uptake. In our study, over half of the participants (52.74%) reported that they chose to get vaccinated to decrease their risk of getting influenza, further reflecting this health awareness.

Broader literature emphasizes the role of social influences in vaccination behaviors (7, 8, 10, 31). The consensus is that social encouragement from family, coworkers and, most notably, healthcare providers significantly promotes vaccination. Influweb participants indicated that they vaccinated due to recommendations from their doctors (17%) or because their school/workplace encouraged it (9%).

Furthermore, 32.55% of participants cited reducing the risk of spreading influenza to others as a motivation, indicating a degree of public health consciousness among the Influweb cohort. A significant percentage (45.49%) also reported that they always get the vaccine, possibly due to established health routines or enhanced access through retirement health plans. This routine behavior is consistent with the generally higher rate of vaccination observed in this population compared to the national average.

The relationship between cumulative incidence of influenza-like illness (ILI) and vaccination rates has been explored previously, with mixed results. Research conducted across 14 European countries found inconsistent correlations between influenza vaccination coverage and ILI incidence, with significant positive correlations observed in some countries but not others (37). In Italy, no significant correlation was found between vaccination coverage and ILI incidence at the national level over the 1999-2000 to 2013-2014 flu seasons (37).

However, our study, which focused on a smaller geographical scale at the macro-regional level, found clearer associations: regions with higher cumulative ILI incidence were significantly linked to increased vaccination rates. This suggests that in macro-regions experiencing higher incidences of ILI, there may be a greater perceived need for vaccination, driving higher uptake among the population.

The COVID-19 pandemic had a significant impact on influenza vaccination behavior, with Influweb participants having 667.5% higher odds to vaccinate during the pandemic. This finding aligns with global studies from Italy (38), England (39), and the United States (40), all of which reported increased flu vaccination rates during the pandemic. The heightened health alert during the pandemic likely contributed to the rise in vaccinations, particularly in health-conscious populations like Influweb’s. Moreover, many individuals may have been offered flu vaccinations alongside COVID-19 vaccinations, further boosting flu vaccine uptake. While some studies in the U.S. suggest that COVID-19 vaccine hesitancy could have a negative effect on flu vaccination uptake (41), our results indicate a strongly positive effect of the pandemic on influenza vaccination rates in Italy.

A study of healthcare workers in Italian cancer centers found a steady increase in influenza vaccination rates between the 2018 and 2021 flu seasons (42). This upward trend mirrors the rise in vaccination coverage observed in the Influweb population, which experienced a notable increase beginning in the 2018-2019 influenza season. The pandemic may have further amplified this existing trend, resulting in the elevated vaccination coverage we observed in this study.

While this study provides valuable insights into vaccination behavior, some limitations should be acknowledged. The discrepancy between Influweb’s self-reported data and national health statistics suggests a potential for reporting biases. First, the use of self-reported data may introduce recall bias, where participants might misremember or misreport their vaccination status. Additionally, the voluntary nature of the Influweb platform may attract participants who are more health-conscious and, as a result, more likely to engage in preventive behaviors such as vaccination. This self-selection bias should be taken into account when interpreting the vaccination rates reported by Influweb participants (43).

Nevertheless, the value of participatory survey platforms should not be underestimated, as they have proven to be reliable tools for disease surveillance, particularly when combined with traditional data sources (19). Future research could benefit from integrating objective vaccination records to validate self-reported data and expanding studies to include more diverse participant populations to ensure broader applicability.

## Conclusions

In conclusion, this study confirms many established determinants of influenza vaccination, such as age, chronic disease, education, and vaccine accessibility, within the Influweb context. It also reveals lower vaccination rates among those living with minors and those who rely on public transportation, despite being at higher risk for ILI infections. The association between higher vaccination uptake during the COVID-19 pandemic and increased cumulative incidence rates of influenza-like illness in macro-regions highlights how health trends can significantly shape individual health behaviors.

Moving forward, leveraging participatory platforms like Influweb could enhance public health efforts by providing real-time insights into population health behaviors, allowing for more responsive and targeted campaigns. Improving accessibility and targeting public health communication efforts at high-risk groups, including public transit users and those who live with minors, could increase vaccination coverage and reduce influenza transmission within these populations.

## Data Availability

The raw data cannot be shared publicly because it contains personal individual information which could compromise user privacy. Data requests should be addressed to the authors.

## Supplementary Materials

### Complete case deletion

Figure S1 compares vaccination coverage rates of the Influweb population before (dark green) and after (purple) the implementation of complete case deletion. Entries with missing values in any of the variables identified by the literature review were excluded from the final dataset. The vaccination coverage rates before and after are highly consistent across all seasons, indicating that the exclusion of data does not significantly bias vaccination coverage rates.

**Figure S1:**
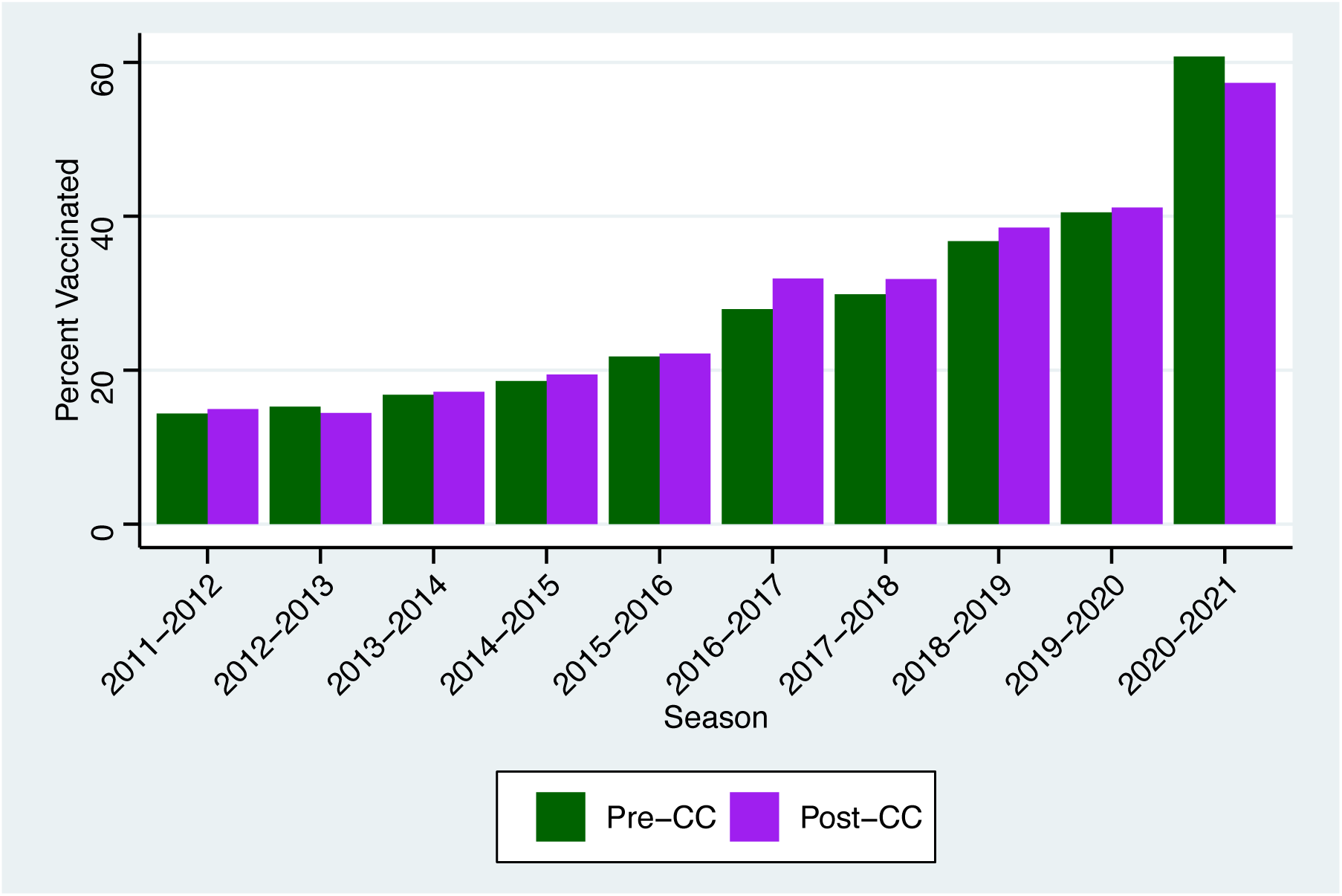
Comparison of vaccination coverage before and after complete case deletion

Table S1 shows the distributional breakdown of important sample demographic factors before and after complete case deletion. This comparison shows that the distribution across age class, sex, macro-region and educational level remain stable. The main source of missing values were the education and macro-region variables. Despite this, the demographic composition of the sample is not distorted after these missing values were excluded.

**Table S1:**
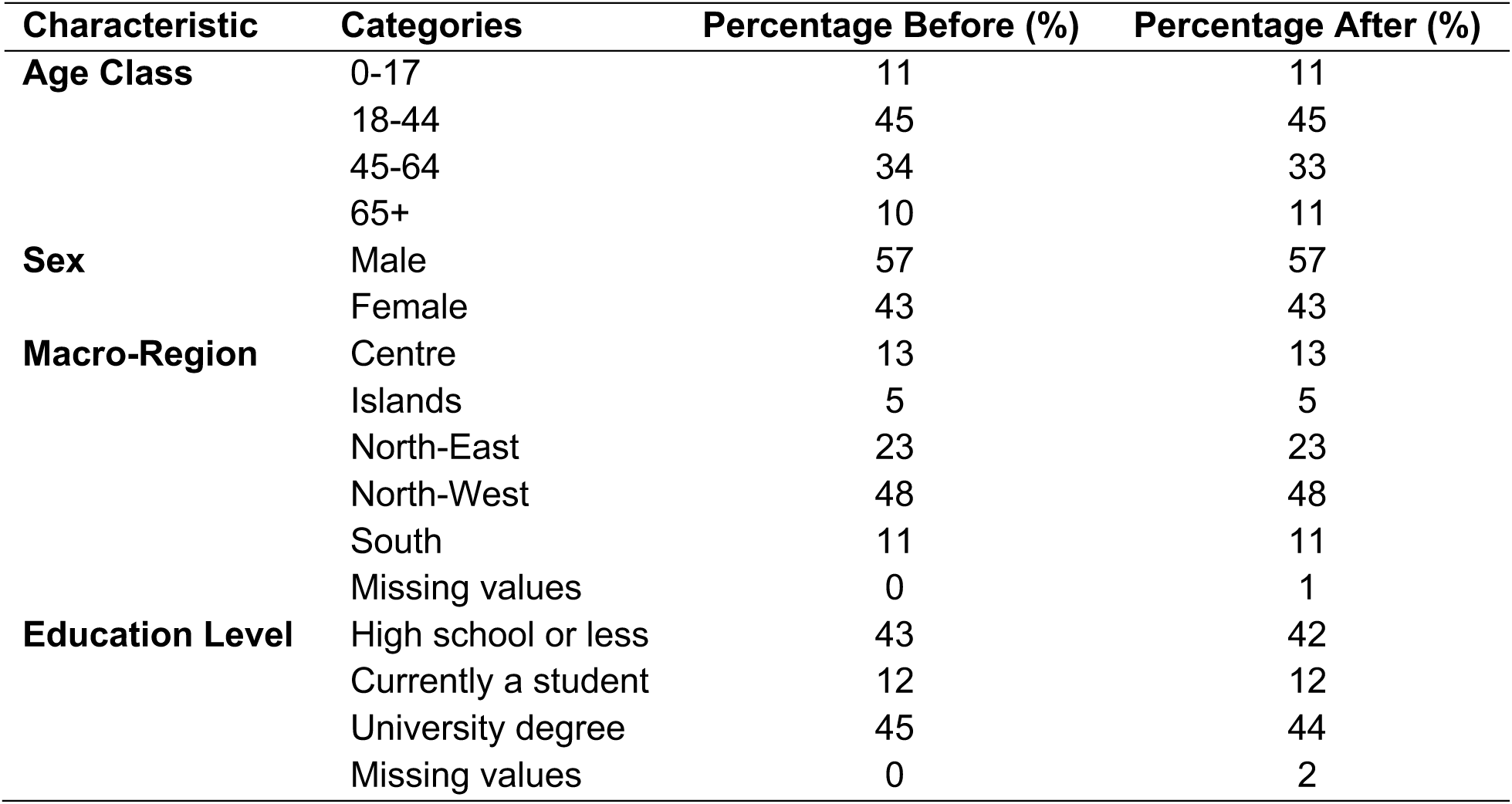
Participant characteristics and their distributions before and after complete-case deletion.

### Retrospective data comparison

To assess the reliability of self-reported vaccination status within the Influweb population, participants were asked whether they were vaccinated in the current flu season and the prior season. For example, during the 2019-2020 season, participants reported whether they were vaccinated in that season as well as in 2018-2019. In this case, we compared the retrospective reports from the 2019-2020 season regarding the 2018-2019 vaccination status with the actual data collected during the 2018-2019 season. This comparison allowed us to validate the consistency and reliability of the self-reported vaccination data over time. Please note that since this was an internal validation analysis, the vaccination rates reported here were not based on weighted data like the other results presented in this study.

**Figure S2:**
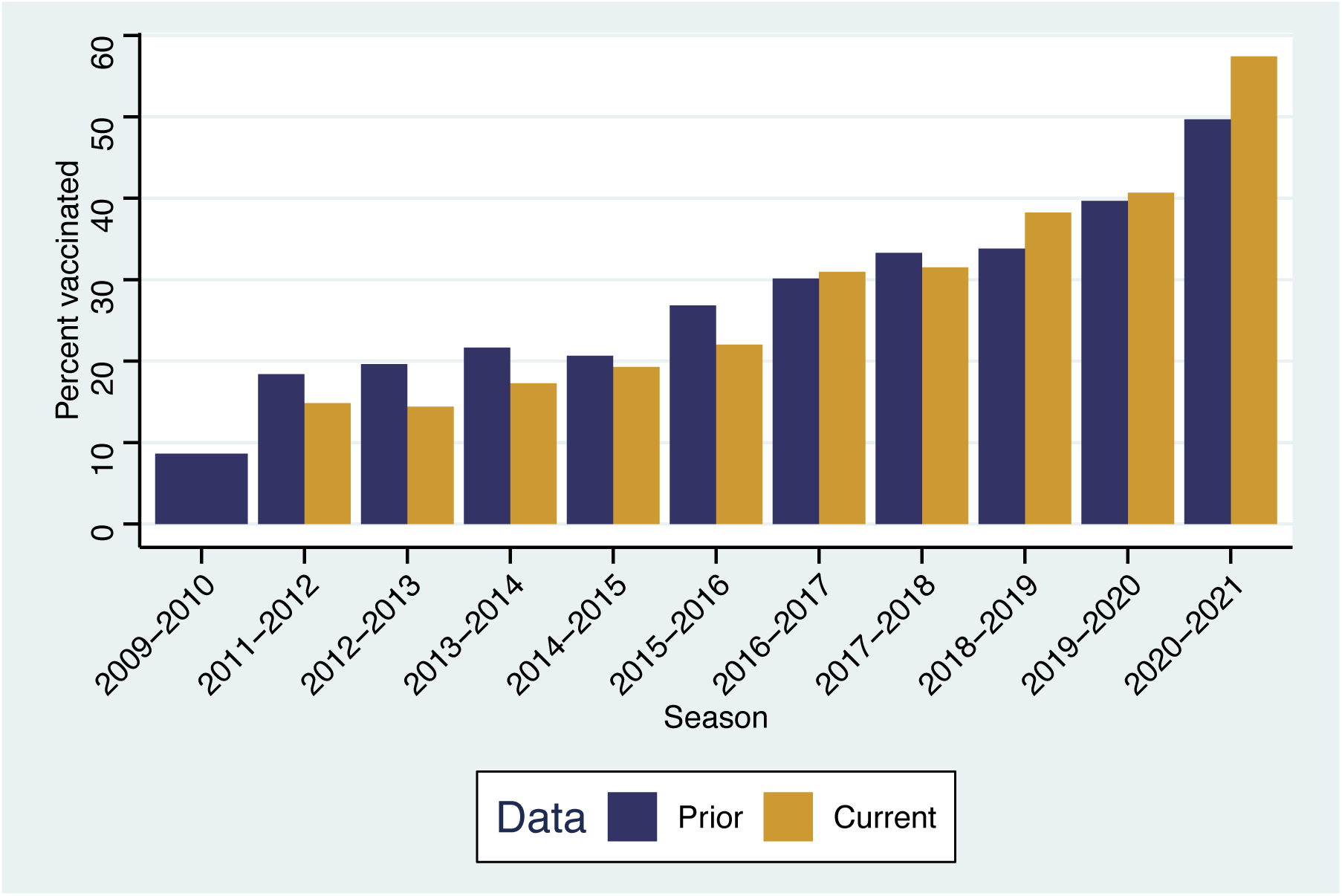
Comparison of self-reported vaccination for the season a survey was submitted and the prior season

Figure S2 illustrates the comparison between self-reported vaccination coverage for the current and prior flu seasons within the Influweb population. Two colors represent the data: the yellow bars indicate the vaccination coverage reported for the current flu season when the survey was conducted, and the dark blue bars represent the coverage reported for the prior flu season. Both follow a similar trajectory with a general upward trend over the years, indicating consistency in the data and an increase in reported vaccination rates over time for both the current and prior seasons.

## Declarations

### Ethics approval and consent to participate

For influweb.it the research was conducted in agreement with Italian regulations on privacy and data collection and treatment. The institutional review board of ISI Foundation, upon consultation with the Italian Data Protection authority, waived the ethical approval for this study.

### Consent for publication

Not applicable.

### Availability of data and materials

Individual data collected for this study cannot be publicly shared due to privacy requirements.

### Authors’ contributions

All authors contributed to the design of the study. DP designed the original survey. Data were processed and prepared for analyses by NG, MM and KK. KK performed the statistical analysis, with oversight from DP, MM and NG. The first draft of the manuscript was produced by KK and all authors reviewed, edited, and approved the final version.

## Acknowledgements

The authors are grateful to Matteo Delfino and Giovanni Frigione for the technical support with the data collection and extraction.

